# Factors Affecting Maternal and Neonatal Mortality in Northern Nigeria: A Multiple Linear Regression Analysis

**DOI:** 10.1101/2022.06.30.22276507

**Authors:** Obinna Orjingene, Ogojah Teryila, Peter Baffoe, Olumuyiwa Ojo

## Abstract

Nigeria has a maternal mortality rate (MMR) of 512 deaths per 100,000 live births, an estimate which indicates that maternal deaths are responsible for about a third of all deaths among women of reproductive age. The regional distribution of neonatal deaths in Nigeria showed that the North East region had the highest neonatal deaths. This study provides insight into identified factors and their influence on maternal and neonatal mortalities. Targeted policy implementation will emerge from the analysis of these factors with the aim of reducing the mortalities which will invariably contribute to the reduction of the global maternal and newborn mortality index

Multiple linear regression analyses using secondary time series data from the district health information system (DHIS2) for the period 2012-2021. Multivariable logistic regression analyses were also used to examine a series of predictor variables to determine those that best predict the outcome variables. Statistical significance for all regressions performed was determined at *p*□<□0.05.

Regression results showed a negative relationship between antenatal care and neonatal mortality implying that an increase in the number of women attending ANC will lead to a reduction in neonatal mortality by about 43%. The regression result showed a positive relationship between low birth weight and neonatal mortality implying that an increase in the number of live births with low birth weight will lead to an increase in neonatal mortality by 94%. Regression analysis on factors affecting maternal mortality showed that 4^th^ antenatal care visits and above, health facility delivery, postnatal care within 3 days for mothers, and skilled birth attendance all have a negative relationship with maternal mortality. The regression results are all statistically significant (p<0.05).

The study revealed significant relationships between some factors (antenatal care, low birth weight, skilled birth attendants, health facility delivery, post-natal care for both mother and newborn) affecting maternal and neonatal mortality.

## 1. Introduction

The poor maternal and neonatal mortality indices in Nigeria have remained a serious public health issue.^1,2^. ^3^Asserted that Nigeria’s neonatal mortality rate is among the worst in the world, Nigeria is also among the top six countries in the world that contribute to more than 50% of all global maternal deaths.^1^

Nigeria has a maternal mortality rate (MMR) of 512 deaths per 100,000 live births,^4^ an estimate which indicates that maternal deaths are responsible for about a third of all deaths among women of reproductive age.^5^ The situation is much worse within the northern parts of the country, where the MMR is estimated to be over 1000 deaths per 100,000 live births.^6 1^ in their study also found a higher maternal mortality rate in Northern Nigeria. Nigeria’s neonatal mortality on the other hand is estimated at 39 deaths per 1000live births.^4^ Nigeria ranks second to India with the highest number of neonatal deaths globally.^7^ According to,^8^ more than 250,000 neonates die every year in Nigeria; this translates to approximately 690 neonates every day. The regional distribution of neonatal deaths in Nigeria showed that the North East region had the highest neonatal deaths.^3^ This is consistent with findings of the National Demographic Health Survey (NDHS) 2018 which also found the Northern region of Nigeria as having the highest burden of neonatal mortality.

The Northern region of Nigeria bears the greater burden of both maternal and neonatal mortality. It is therefore pertinent to identify and analyze factors that are responsible for the high burden of these mortalities. This would provide more insight into the factors and their influence on maternal and neonatal mortalities. Targeted policy implementation will emerge from the analysis of these factors with the aim of reducing the mortalities which will invariably contribute to the reduction of the global mortality index.

## 2. Maternal mortality and its causes

Maternal death or mortality is defined as the death of a woman while pregnant or within 42 days of pregnancy, expressed as a ratio to 100,000 live births in the population being studied.^9^

According to,^10^ about 80% of maternal deaths globally are due to four major causes-severe bleeding, infections, hypertensive disorders in pregnancy (eclampsia), and obstructed labor. ^11^ estimated that in Nigeria, more than 70% of maternal deaths could be attributed to five major complications: hemorrhage, infection, unsafe abortion, hypertensive disease of pregnancy, and obstructed labor.

The causes of maternal deaths can be classified into medical factors, health factors, reproductive factors, unwanted pregnancy, and socioeconomic factors.^12^ Medical factors include direct obstetric deaths, indirect obstetric deaths, and unrelated deaths. Direct obstetric deaths result from complications of pregnancy, delivery, or their management. Indirect obstetric deaths result from worsening of some existing conditions (such as hepatitis) by pregnancy. Health service factors include deficient medical treatment, mistaken or inadequate action by medical personnel, lack of essential supplies and trained personnel in medical facilities, lack of access to maternity services, and lack of prenatal care. Other risk factors for maternal mortality in Nigeria include maternal age, illiteracy, non-utilization of antenatal services, and grand multi-parity.^13^

### 2.2 Neonatal mortality and its causes

Neonatal mortality refers to the incident of death occurring within the first 28□days of life. Causes of neonatal mortality have been identified by several studies. A study by ^2^ identified low birth weight, lack of antenatal care, maternal illness, mother’s age, prematurity, and birth asphyxia as causes of neonatal mortality. ^14^ in their study found severe perinatal asphyxia, low birth weight, and infections as leading causes of neonatal deaths.

Findings of the 2019 Verbal and Social Autopsy Study (VASA) revealed that the leading causes of neonatal death are sepsis, intrapartum injuries, and pneumonia, with the majority of neonatal deaths occurring in the early neonatal period (first seven days). The physician coding finds jaundice and preterm birth as important causes of death (9-10% of cases each), but these are uncommon in the expert algorithm (1% each). Both methods find that congenital abnormalities, diarrhea, injuries, and neonatal tetanus, while present, cause only a few percent of deaths in neonates.

In a study by,^15^ it was revealed that preterm babies’ risk of death is 12 times higher than that of full-term babies and has an increased risk of disability. Even though prematurity is principally an influencing condition, which occurs in severe immaturity, death, if occurred, is a result of complications that account for about one-third of all neonatal deaths.^15^ As documented by USAID, adverse intrapartum events (birth asphyxia) account for about 31 percent of neonatal deaths in Nigeria. The cause is very much associated with quality of care during childbirth. Infections (sepsis, pneumonia, tetanus, and meningitis) result in over 26% of neonatal deaths.^16^ Some complications of prematurity as highlighted are also related to infections that may cause neonatal mortality to a greater extent. A social and verbal autopsy report carried out in Nigeria in 2014 by USAID, revealed sepsis as the leading (31.5%) cause of neonatal death.

Other causes of mortality in newborns in Nigeria include poor quality of care. Health services are provided through both public and private sectors with primary healthcare being a primary significant. But, the accessibility of these services does not equate to good quality of care. Private health care service is poorly incorporated into Nigeria’s health system even though it plays a significant role in rending care. Other challenges to optimal health care services include the distance to be covered to reach health facilities, especially in rural areas, the cost of services, disruption of services, poor quality of care, inadequate implementation of the standard guidelines, and attitudes of health workers to care of patients.^17^ There are intense disparities in coverage and quality of care at birth, where the majority of births take place in a health facility and with a skilled attendant but still, the quality of care remains low with poor outcomes for mothers and babies. According to Demographic Health Survey, 2018, rural and less educated women are less likely than others to attend ANC, have assistance from a skilled health provider during delivery, and give birth in a health facility.^4^ Infection during and after childbirth is high in Nigeria as a result of the high occurrence of home births; therefore, prevention is extremely important. In view of this, FGON provides through the FMOH clean home delivery kits (Mama Kits), and in addition, the use of 4% chlorhexidine gel has been approved for cord care by the FMOH at the community as well as facility levels, and implementation is planned at scale.^18^

### 2.3 Strategies to improve maternal and newborn health and reduce neonatal deaths in Nigeria

^19^ Reported strategies that were effective in reducing maternal mortalities; community-based maternal and child health antenatal care focused strategies, emergency transport scheme, community-based distribution of misoprostol tablets to mothers in the third stage of labor for the prevention of postpartum hemorrhage, etcetera. Several strategies have been put in place by both the federal and state governments and partners to reduce maternal and neonatal deaths in the country.

**Table 1.**

| S/N | Key Policy and Strategies documents | Goal | Description |
| --- | --- | --- | --- |
| 1 | National Strategic Health Development plan (NSHDP) II | The NSHDP II addresses lingering and emerging health sector challenges. It also offers the opportunity to ensure better health outcomes by 2022 through consolidation of the gains made and incorporation of the lessons learned from NSHDP I. | Some of the challenges identified in the NSHDP I end-term evaluation and which have been considered in NSHDP II include:<br>*gaps in political will and poor program ownership at lower levels especially state and LGA levels;<br>*weak donor coordination and harmonization of development and technical assistance;<br>*low level of government financing of healthcare at the three levels of government;<br>*weak M&E systems to monitor implementation of the State strategic Health Development Plans and<br>* weak Primary Health Care structures. |
| 2 | Child Health Policy (Revised 2018). | The Child Health Policy document will serve to provide a platform for the systematic development and implementation of evidence-based interventions for improved child health in Nigeria. | The policy proposes strategies and actions that aim to reduce the unacceptably high rates of morbidity and mortality of children in Nigeria, securing a better future for them |
| 3 | Reproductive, Maternal, Newborn, Child, Adolescent Health + Nutrition Strategy for Nigeria: 2007 | The IMNCH Strategy aimed at fast-tracking coverage of effective interventions along the continuum of care towards the reduction of maternal, neonatal and child morbidity and mortality in line with Millennium Development Goal (MDG) 4 and 5 targets. | This strategy document is being proposed for review, in order to reflect the SDG target for NMR of 12/1000 live birth by 2030. |
| 4 | National guidelines on Maternal, Perinatal, and Child Death Surveillance Response (MPCDSR) | The prompt response to the recommendations made during the audits of the maternal, perinatal and child deaths will improve quality of care, reduce maternal, newborn and child deaths significantly in Nigeria. | The MPCDSR guideline and the tools provide direction and instructions required for the establishment of Maternal, Perinatal and Child Death Surveillance Response in Nigeria. |
| 5 | Essential Newborn Care Course | It aims to ensure health workers have the skills and knowledge to provide appropriate care at the most vulnerable period in a baby's life | The Essential Newborn Care Course (ENCC) was adopted in 2008 and harmonized in 2016. |
| 6 | Community Based Newborn Care (CBNC) Course | which is aimed at increasing the coverage of household and community interventions that will reduce newborn and child mortality and promote the healthy growth and development of young children. | In 2011, Nigeria adopted the CBNC course, which is part of the WHO-UNICEF package "Caring for Newborns and Children in the Community" The package consists of 3 courses, namely on caring for the newborn at home, promoting healthy growth and development, and caring for the sick child. |
| 7 | National Strategy for Scale-up of Chlorhexidine in Nigeria | This strategy seeks not only to integrate Chlorhexidine into existing health programs but also to strengthen the underlying systems that will support the scale-up of other products for mothers and children in Nigeria | The strategy proposes concrete interventions across five core components of scale-up: market & user; manufacturing & distribution; clinical & regulatory; policy, advocacy, & financing; and coordination. |
| 8 | Nigeria Every Newborn Action Plan (NiENAP) | The aim was the plan to serve as a framework for each of the 36 state and Federal Capital Territory (FCT) and collaborate with many stakeholders and partners to develop their own action plans | In 2017, the Federal Ministry of Health launched Nigeria Every Newborn Action Plan to end preventable deaths in the country. |
| 9 | Nigeria Essential Medicine List 2020 |  | The seventh edition of the Nigeria Essential Medicines List (NEML), is a veritable tool in the healthcare delivery system which forms the fulcrum for the treatment of various medical conditions prevalent in the country, especially for newborns. |
| 10 | National Reproductive Health Policy (2010-2015) | 75% reduction in maternal mortality from 1000/100,000 in 1990 to 250/100,000 live births by 2015 (in line with MDG5.) | • Increase the proportion of pregnant women delivered by skilled attendants from 37 percent in 2008 to 78 percent by 2015.<br>• Increase the proportion of births taking place in facilities meeting the essential obstetric standard from 4 percent in 2002 to at least 15 percent by |
|  |  |  | 2015. |
| 11 | National Acceleration Plan for Pediatrics and Adolescent HIV Treatment and Care (2020-2022) | Identifies the main factors that limit access to HIV services along the entire cascade for children and adolescents and recommends practical strategies for addressing them. | A road map for the unmet needs of the estimated 140,000 and 121, 000 children and adolescents respectively living with HIV in Nigeria accessed ART in 2018 |
| 12 | Task Shifting and Task Sharing Policy for Maternal and Newborn Health Care in Nigeria | To provide a legal framework for empowering community health workers (CHOs and CHEWs) to provide quality maternal and newborn care services, especially at PHCs | Addresses the human resource shortages that militate the provision of critical Maternal and Newborn services |
| 13 | Accelerated Reduction of Maternal and Newborn Mortality in Nigeria | Identifies the Road Map for Accelerated Reduction of Maternal and Neonatal Mortality to guide implementation especially in the high burden States | Clear cut measures to guide implementers and stakeholders at all levels to crystallize results on ending preventable maternal and newborn deaths |
| 14 | National Integrated Pneumonia Control Strategy and Implementation Plan (2019) | The plan promotes an integrated approach to pneumonia control through multi-sectoral actions | Pneumonia control is central to achieving Universal Health Coverage and meeting Sustainable Development Goals in Nigeria |
| 15 | Coordination Platforms | Facilitates synergy in the implementation of Maternal and newborn health programs in the Country. | Different Coordination fora at the national and sub-national level; Human Resource for Health, Quality of Care, Child Health, Reproductive Health Technical Working |
|  |  |  | Groups, and Newborn subcommittee. |

## 3. Methodology

Secondary time series data for this study were retrieved from the district health information system (DHIS2) for the period 2012-2021, a 10-observation data point per variable. An appropriate number of observations can produce accurate results. Moreover, the results from a small number of observations will produce questionable results. There is however no certain rule of thumb to determine the number of observations. For example, in regression analysis, many researchers say that there should be at least 10 observations per variable. If we are using three independent variables, then a clear rule would be to have a minimum sample size of 30.

### 3.1 Variables

Two dependent or outcome variables were used in the study; maternal deaths, and neonatal deaths. The factors considered in this study to influence maternal deaths (independent variables) were; Ante-natal care attendance up to 4^th^ visits, health facility delivery, post-natal care attendance within three days for mothers, and skill birth attendance. On the other hand, factors considered in this study to influence neonatal deaths (independent variables) were; low birth weight, skill birth attendance, post-natal care checks for newborns, ante-natal care attendance, and facility delivery.

### 3.2 Statistical analyses

Statistical analyses were performed using SPSS22. Trend analyses for maternal and neonatal deaths as well as factors that influence them were conducted. Multivariable logistic regression analyses were also used to examine a series of predictor variables to determine those that best predict the outcome variables. Statistical significance for all regressions performed was determined at *p*□<□0.05. In addition to trend and regression analyses, Spearman rank correlation analysis was performed to examine the linear association between the independent variables.

The regression equation to examine factors affecting neonatal mortality is given below

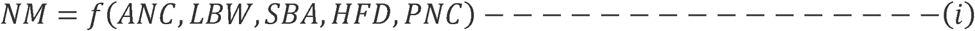

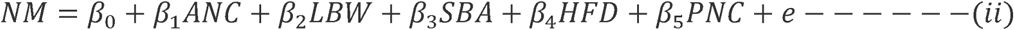

Where:

NM=Neonatal mortality

ANC=Ante-natal care attendance

LBW=Low birth weight

HFD=Health facility delivery

PNC=post-natal care within 3days-newborn

SBA=skilled birth attendance

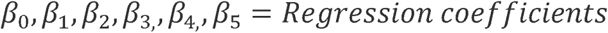

e=error term

Similarly,

The regression equation to examine factors affecting maternal mortality is given below

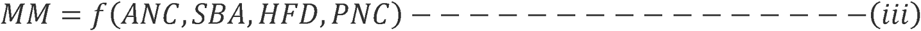

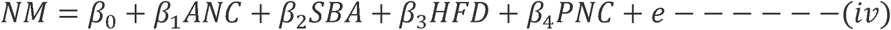

Where:

MM=Maternal mortality

ANC=Ante-natal care attendance

HFD=Health facility delivery

PNC=post-natal care within 3days-mother

SBA=skilled birth attendance

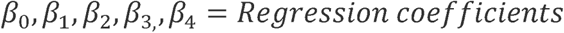

e=error term

The Spearman rank correlation is computed using the formula

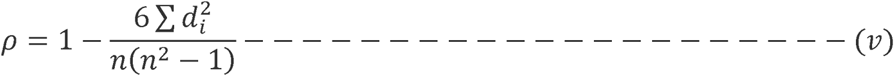

Where: *ρ* = Spearman rank correlation

*d* = *difference between the two ranks of each observation*

*n* = *number of observations*

### 3.3 Assumptions

The data used in the study are derived from a random, or at least representative, sample of the population. Study variables are jointly normally distributed random variables. They follow a bivariate normal distribution in the population from which they were sampled.

### 3.3 Limitation

In this study, however, there is a limited number of yearly observations both on the DHIS2 and from available survey reports (NDHS, MICS).

## 4. Results

Maternal mortality declined between 2013 and 2018 from 576 to 512 deaths per 100,000 live births. The neonatal mortality trend on the other hand showed an increase in neonatal deaths between the same period from 37 to 39 deaths per 1000 live births.

The maternal mortality ratio for the 19 Northern States was calculated using data from dhis2. The trend for a 5-year period is depicted in the figure 3 below.

**Fig 1:**
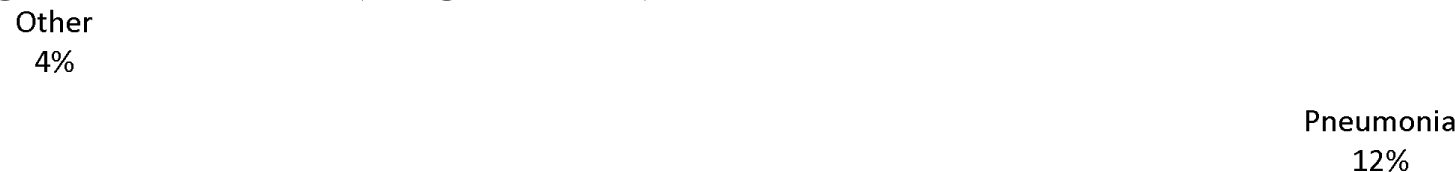
Physician-coded and expert algorithm verbal autopsy for causes of 722 neonatal (0-27 days) deaths in Nigeria, 2013-2018 (weighted data)

**Fig 2.** Trends in maternal mortality rates

**Fig 3.** Trends of maternal mortality rate in Northern Nigeria (Dhis2 2017-2021).

Similarly, the neonatal mortality ratio for the 19 Northern States was calculated using data from dhis2. The trend for the 10-year period is depicted in the figure 4 below.

**Fig 4.** Trends in neonatal mortality rates

**Fig 5.** Trend of neonatal mortality rate in Northern Nigeria.

### 4.1 Regression analysis

#### Regression analysis on factors affecting neonatal mortality

| Model |  | Unstandardized Coefficients |  | Standardized Coefficients | t | Sig. |
| --- | --- | --- | --- | --- | --- | --- |
|  |  | B | Std. Error | Beta |  |  |
| 1 | (Constant) | .894 | .000 | .856 | .436 | .000 |
|  | ANC | -.437 | .000 | -.430 | .219 | .000 |
|  | LBW | .968 | .000 | .947 | .295 | .000 |
|  | HFD | -.785 | .000 | -.779 | .389 | .000 |
|  | PNC-c | -.852 | .000 | -.844 | .461 | .000 |
|  | SBA | -.679 | .000 | -.618 | .321 | .000 |

a. Dependent Variable: NM

Regression results showed a negative relationship between antenatal care and neonatal mortality implying that an increase in the number of women attending ANC will lead to a reduction in neonatal mortality by about 43%. The regression result showed a positive relationship between Low birth weight and neonatal mortality implying that increase in the number of live births with low birth weight will lead to increase in neonatal mortality by 94%. This means that newborns with low birth weight have little probability of survival. Health facility delivery and postnatal care within 3 days for newborn both have negative relationship with neonatal deaths implying that increase in these variables will lead to decrease in neonatal deaths by about 78% and 84% respectively. Skilled birth attendants had an inverse relationship with neonatal mortality. This implies that increase in deliveries taken by skilled personnel leads to decrease in number of newborn deaths. The regression results are all statistically significant (p<0.05).

#### Regression analysis on factors affecting maternal mortality

| Model |  | Unstandardized Coefficients |  | Standardized Coefficients | t | Sig. |
| --- | --- | --- | --- | --- | --- | --- |
|  |  | B | Std. Error | Beta |  |  |
| 1 | (Constant) | .914 | .000 | .893 | .393 | .000 |
|  | ANC | -.721 | .000 | -.711 | .431 | .000 |
|  | HFD | -.836 | .000 | -.819 | .349 | .000 |
|  | PNC-m | -.894 | .000 | -.872 | .281 | .000 |
|  | SBA | -.913 | .000 | -.898 | .349 | .000 |

a. Dependent Variable: MM

Regression analysis on factors affecting maternal mortality showed that ante-natal care visits (4^th^ visits and above), health facility delivery, postnatal care within 3 days for mothers, and skilled birth attendance all have negative relationship with maternal mortality. This implies that when all these variables increase, maternal mortality decreases by varying percentages as shown in the result.

### Spearman correlation between independent variables (factors influencing maternal and neonatal mortality)

Strong positive correlation exists between ante-natal care 4th visits and health facility delivery, skilled birth attendance, post-natal care for both mother and child. A moderate correlation was however found between ANC 4th visit and low birth weight. Health facility delivery had a strong positive correlation with post-natal care for both mother and newborn as well as skill birth attendance.

|  |  | ANC | HFD | SBA | PNC-m | PNC-c | LBW |
| --- | --- | --- | --- | --- | --- | --- | --- |
| Spearman<br>Correlation | ANC | 1.000 | .987 | .954 | .963 | .970 | .599 |
|  | HFD | .987 | 1.000 | .999 | .998 | .985 | .247 |
|  | SBA | .954 | .999 | 1.000 | .996 | .999 | .253 |
|  | PNC-m | .963 | .998 | .996 | 1.000 | .995 | .391 |
|  | PNC-c | .970 | .985 | .999 | .995 | 1.000 | .282 |
|  | LBW | .599 | .247 | .253 | .391 | .282 | 1.000 |
| Sig. (1-tailed) | ANC | . | .004 | .002 | .005 | .003 | .019 |
|  | HFD | .004 | . | .000 | .000 | .000 | .058 |
|  | SBA | .002 | .000 | . | .000 | .000 | .280 |
|  | PNC-m | .005 | .000 | .000 | . | .001 | .258 |
|  | PNC-c | .003 | .000 | .000 | .001 | . | .024 |
|  | LBW | .019 | .058 | .280 | .258 | .024 | . |
| N | ANC | 10 | 10 | 10 | 10 | 10 | 10 |
|  | HFD | 10 | 10 | 10 | 10 | 10 | 10 |
|  | SBA | 10 | 10 | 10 | 10 | 10 | 10 |
|  | PNC-m | 10 | 10 | 10 | 10 | 10 | 10 |
|  | PNC-c | 10 | 10 | 10 | 10 | 10 | 10 |
|  | LBW | 10 | 10 | 10 | 10 | 10 | 10 |

## 5. Discussions

Nigeria is the most populous country in Africa and the seventh most populous in the world with an estimated population of 214.4 million in 2020.^20^ Newborn deaths are a significant contributor to Under-five Mortality Rate, it accounts for 45% of U5MR globally and 32% in Nigeria.^21^ The neonatal mortality rate had increased from 37 to 39 per 1000 live birth (NDHS 2018). Maternal mortality is also high in the country, especially in the northern region with over 1000 maternal deaths per 100,000 live births. The country’s failure to reduce both maternal and neonatal Mortality Rates will hamper the attainment of the SDG 3 targets.

The analysis of ten-year data has brought to the limelight the likely significant contribution of factors such as ante-natal care attendance, health facility delivery, postnatal care for both mother and newborn, low birth weight, and skilled birth attendance on maternal and neonatal mortality. Results revealed that increased uptake of ANC services will save 43% of deaths of newborns in Northern Nigeria. The inverse relationship between ANC and both maternal and neonatal deaths shown in this study is consistent with other studies. ^2,22,23,24,1^ Though these studies were conducted at different times results are still consistent with the present study. This is evident that ANC is an important factor in the reduction of both maternal and neonatal mortalities. Antenatal care is a major component of reproductive health care and consists of prenatal, natal, and postnatal care which aims at reducing infant and maternal morbidity and mortality through early detection of complications and prompt treatment, prevention of diseases, birth preparedness and health promotion. In addition, ANC period is an opportune time for reaching pregnant women with a number of additional interventions that may be vital to the health and well-being as well as to the health of their unborn children. It involves the percentage increase or frequency in the number of visits for antenatal care and delivery place. It also promotes hospital or facility-based continuum of care during pregnancy and childbirth as shown in the correlation analysis. The findings also showed that newborns with low birth weight had low probability of survival. This is in line with findings of a study by, ^25^ who also found birth weight as a significant predictor of neonatal mortality. Findings of this study showed that deliveries conducted by skilled personnel contributed to reduction of neonatal deaths. This finding is consistent with that of.^23^ This is obvious as skilled personnel are trained to effectively handle complications that may arise during or after labor which may lead to the death of the mother and /or newborn.

## 6. CONCLUSION AND RECOMMENDATIONS

### 6.1 Conclusion

Independent variables that influence simultaneously or partially on maternal and neonatal Mortality Rate are ANC completion, low birth weight, and skilled birth attendance with low birth weight having the greatest influence on neonatal deaths. The study revealed significant relationships between some factors (antenatal care, low birth weight, skilled birth attendants, health facility delivery, post-natal care for both mother and newborn) affecting maternal and neonatal mortality. ANC visits and skilled birth attendants have been proven by the study to be effective in reducing maternal and neonatal deaths. The study also showed low birth weight as the major contributor to neonatal deaths.

### 6.2 Recommendations

To accelerate the survival of neonates in the study area, the study recommends

- An integrated approach ensuring continuity of care through pregnancy, delivery and postpartum period is essential.
- Government should ensure plans to improves healthcare services for maternal and new-born by equipping health facilities with adequate facilities, commodities, and manpower for effective maternal and newborn care service delivery.
- The Nigeria Every Newborn Action Plan should be domesticated across the 19 states and the state should be supported to fully operationalize the plan

## CONSENT

It is not applicable.

## Supporting information

Ethics Committee Waiver letter

## Data Availability

The data set used for the analysis have been submitted as a supporting information

## ETHICAL APPROVAL

It is not applicable, as only secondary data was used for the analysis.

## COMPETING INTERESTS

Authors have declared that no competing interest exist.

## Notes

### Competing Interest Statement

The authors have declared no competing interest.

### Funding Statement

This article did not receive any funding from any source

### Author Declarations

Ethical oversight was waived by the National Health Research Ethics committee of Nigeria (NHREC), please attached waiver letter in supplementary files.

