## Supplementary material for "Factors Affecting Maternal and Neonatal Mortality in Northern Nigeria: A Multiple Linear Regression Analysis": Ethics Committee Waiver letter

**National Health Research Ethics Committee of Nigeria (NHREC)**

Promoting Highest Ethical and Scientific Standards for Health Research in Nigeria

Federal Ministry of Health

**NHREC Protocol Number NHREC/01/01/2007-17/06/2022**
**NHREC Approval Number NHREC/01/01/2007-28/06/2022**
**Date: 28th June, 2022**

**Re: Factors Affecting Maternal and Neonatal Mortality in Northern Nigeria: A Multiple Linear Regression Analysis**

Health Research Ethics Committee (HREC) assigned number: NHREC/01/01/2007

Name of Principal Investigator: Dr Obinna Orjingeno

Address of Principal Investigator: RMNCH Technical Advisor
John Hopkins Program for International Education in Gynecology& Obstetric
JHPIEGO Nigeria Country office


Date of receipt of valid application: 17/06/2022

Date when final determination of research was made: 28-06-2022

**Notice of Research Exemption**

This is to inform you that the activity described in the submitted protocol/documents have been reviewed and the Health Research Ethics Committee has determined that according to the National Code for Health Research Ethics, the activity described there-in meets the criteria for exemption and is therefore approved as exempt from NHREC oversight.

The National Code for Health Research Ethics requires you to comply with all institutional guidelines, rules and regulations and with the tenets of the Code. NHREC reserves the right to conduct compliance visit to your research site without previous notification.

Signed

**Professor Zubairu Iliyasu MBBS (UniMaid), MPH (Glasg.), PhD (Shef.), FWACP, FMCPH, FFPH(UK)**
**Chairman, National Health Research Ethics Committee of Nigeria (NHREC)**

Department of Health Planning, Research & Statistics
Federal Ministry of Health
11th Floor, Federal Secretariat Complex Phase III
Ahmadu Bello Way, Abuja

,
URL: http://www.nhrec.net,
